# A study of SARS-CoV-2 epidemiology in Italy: from early days to secondary effects after social distancing

**DOI:** 10.1101/2020.04.06.20055392

**Authors:** Marco Claudio Traini, Carla Caponi, Riccardo Ferrari, Giuseppe Vittorio De Socio

## Abstract

**Background:** The outbreak of severe acute respiratory syndrome coronavirus 2 (SARS-CoV-2) has led to 101739 confirmed cases, in Italy, as of March 30th, 2020. While the analogous event in China appears to be under control at the moment, the outbreaks in western countries are still at an early stage of development. Italy, at present, is playing a major role in understanding the transmission dynamics of these new infections and evaluating the effectiveness of control measures in a western social context.

**Methods:** We combined a quarantined model with early-stage development data in Italy (during the period February 20th - March 30th) to predict longer-term progression (up to March 30th, and till June 25th, 2020 in a longterm view) with different control measures. Due to significant variations in the control strategies, which have been changing over time, and thanks to the introduction of detection technologies leading to faster confirmation of the SARS-CoV-2 infections, we made use of time-dependent contact and diagnosis rates to estimate when the effective daily reproduction ratio can fall below 1. Within the same framework, we analyze the possible secondary infection event after relaxing the isolation measures.

**Outcomes and interpretation:** We study two simplified scenarios compatible with the observation data and the effects of two stringent measures on the evolution of the epidemic. On one side, the contact rate must be kept as low as possible, but it is also clear that, in a modern developed country, it cannot fall under certain minimum levels and for a long time. The complementary parameter tuned is the transition rate of the symptomatic infected individuals to the quarantined class, a parameter *δ*_*I*_ connected with the time *t*_*I*_ = 1*/δ*_*I*_ needed to perform diagnostic tests. Within the conditions of the outbreak in Italy, this time must fall under 12-8 hours in order to make the reproduction number less than 1 to minimize the case numbers. Moreover, we show how the same parameter plays an even more important role in mitigating the effects of a possible secondary infection event.

**Funding:** No funding has been provided by private or public actors beyond the current clinical practice (National Health Service).

## I. INTRODUCTION

The outbreak of the severe acute respiratory syndrome coronaviruses (SARS-CoV-2) [1–3] has been defined pandemic on March 11th, 2020 due to the global spread. The respiratory droplet transmission is the main contagious route of SARS-CoV-2, and it can also be transmitted through contact [4]. The latency period is generally from 3 to 7 days, with a maximum of 14 days [5], and unlike SARS-CoV, SARS-CoV-2 is contagious during the latency period [6, 7]. In Europe, Italy is becoming a particularly alarming and interesting place to study the evolution of the epidemic, also thanks to the detailed information offered by the Italian Health organizations and the relevant control measures adopted to prevent transmission. Based on Chinese experience and the estimation of transmission models published by Tang *et al*. [8, 9], we develop an approach to the evolution of the SARS-CoV-2 during the early stages of transmission in Italy (see also ref. [10]). Our findings may be useful for inference, forecasting, or scenario analysis. Even though the epidemic is changing rapidly, and our results have to be considered an estimate, the models we are using can be considered predictive and useful for the interpretation of such an unexpected event. A key factor strongly influencing the SARS CoV-2 outbreak progression is the (effective) viral reproduction number (*R*_0_) defined as the mean number of secondary cases generated by a typical infectious individual in a fully susceptible population, the mean *R*_0_ range was estimated from 2.24 (95%CI: 1.962.55) to 3.58 (95%CI: 2.894.39) in the early phase of the outbreak in Cina [11]. We aim to study the relevant parameters of control strategies that lower the reproduction rate of SARS-CoV-2 and mitigate the consequences of the restoration of social normalcy. This last aspect is studied in some detail to investigate the measures to be taken to start restoration without running into significant secondary events [12].

## II. METHODS AND ANALYSIS

The model we use in order to parametrize the Italian outbreak is the generalized *SEIR*-type epidemiological employed model by Tang *et al*. [8]. This model incorporates appropriate compartments relevant to intervention such as quarantine, isolation, and treatment. The population is stratified in Susceptible (*S*), exposed (*E*), asymptomatic infected individuals (*A*), infectious with symptoms (*I*), hospitalized (*H*), and recovered (*R*). Further stratification includes quarantined susceptible (*S*_*q*_), and isolated exposed (*E*_*q*_) compartments (see Fig. 1).

**FIG 1.**
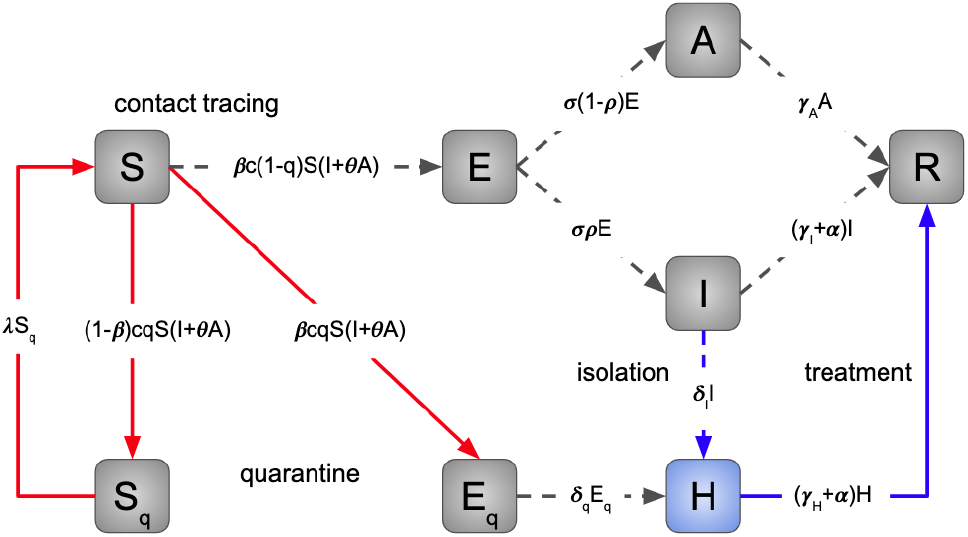
Diagram of the model simulating the novel Coronavirus (Sars-CoV-2) infection in Italy. The population is stratified in Susceptible (*S*), exposed (*E*), asymptomatic infected individuals (*A*), infectious with symptoms (*I*), hospitalized (*H*) and recovered (*R*), quarantined susceptible (*S*_*q*_), isolated exposed (*E*_*q*_) compartments. Interventions like intensive contact tracing followed by quarantine and isolation are indicated (cfr. ref. [8]).

A portion of susceptibles, *S*, get in contact with infected individuals with rate *c* (*I* + *θA*), where *c* is the contact rate, *I* the number of symptomatic infected individuals, *A* the asymptomatic infected individuals, and *θ <* 1 the contribution of asymptomatic infected to the infection spread. With contact tracing, a proportion, *q*, of susceptible, *S*, that get in contact with infected individuals is quarantined. Individuals receive the virus at rate *β*, which is the transmission probability, and become exposed. On the other side, the exposed individuals identified with contact tracing get quarantined at rate *q*. Therefore, we have three fluxes of individuals out of *S*: the quarantined with virus transmission going into *E*_*q*_, *βcqS*(*I* + *θA*); the quarantined without virus transmission going into *S*_*q*_, (1 *− β*)*cqS*(*I* + *θA*); the individuals with virus transmission but not identified and not quarantined going into *E, βc*(1*− q*)*S*(*I* + *θA*). Quarantined individuals without virus transmission are released at rate *λ*, generating an inbound flux of individuals to *S* given by *λS*_*q*_. Exposed, infected, and quarantined individuals move to the hospitalized compartment at rate *δ*_*q*_*E*_*q*_. Exposed, infected, and not quarantined individuals become infectious at rate *σ*. Some of them develop symptoms with a probability of *ρ*. Then, there are two outbound fluxes of individuals for the compartment *E*: the infected with symptoms, *σρE*; the infected asymptomatic *σ*(1 − *ρ*)*E*. The infected with symptoms will eventually be detected and hospitalized with a rate of *δ*_*I*_, which reflects the sanitary system’s diagnostic capability. Finally, all the infected will recover with rate: *γ*_*A*_, for the asymptomatic, *γ*_*I*_ for the infected not hospitalized, *γ*_*H*_ for the infected hospitalized. The infected with symptoms, both hospitalized or not, have a mortality rate of *α*. For disease transmission, these individuals pass to the recovered compartment, as they are no more infectious. Summing the inbound and outbound fluxes at each compartment, we obtain the system of differential equations of the model:

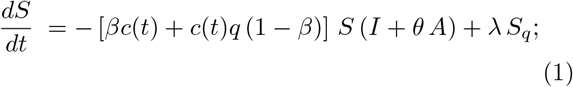

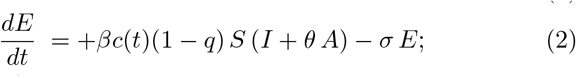

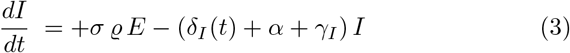

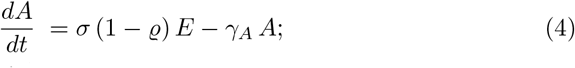

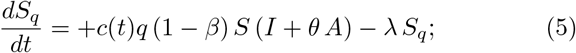

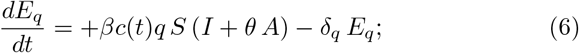

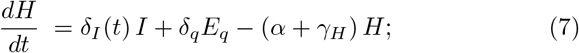

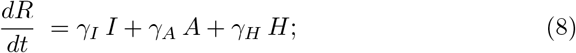

and the values of the parameters are reported in the appendix and discussed in the next Section

### A. A time dependent quarantined model with isolation

On March 8th, 2020, the Italian Government announced the implementation of restrictions for controlling the infection. Many cities and provinces (of different sizes) applied the proposed measures. Control measures have been adopted, like convincing all the residents to stay home and avoid contacts as much as possible. From the mathematical perspective, this can significantly contribute to decreasing the contact rate of *c* among the persons. Besides, the 2019-nCoV tests were gradually shortening the time of diagnosis (i.e., the value of *δ*_*I*_ increases continuously). Considering these control strategies, we could tune the model on the concrete Italian conditions [13].

The equations of the model, shown in Eqs. (1)-(8), contain parameters explicitly dependent on time. In particular the contact parameter *c* and the transition rate of declared infected individuals *δ*_*I*_ (cfr. refs. [8, 9]).

The time-dependence is parametrized as

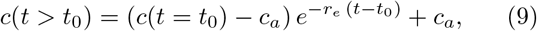

where *c*_*a*_*/c*_0_ = 0.2, *t* = 0 is fixed at February 20th and *t*_0_ selects the initial day of a rapid implementation of the isolation measures for the entire population. At the same time the parameter *δ*_*I*_ fixing the transition rate to quarantine of the infected individuals (*t*_*I*_ = 1*/δ*_*I*_) increases because of a decreasing of the testing time:

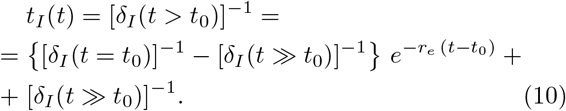

In Fig. 2 the behavior of the two parameters as a function of time. The rate of change *r*_*e*_ = 0.33 day^*−*1^ assumes the same values for the gradually changing quantities.

**FIG 2.**
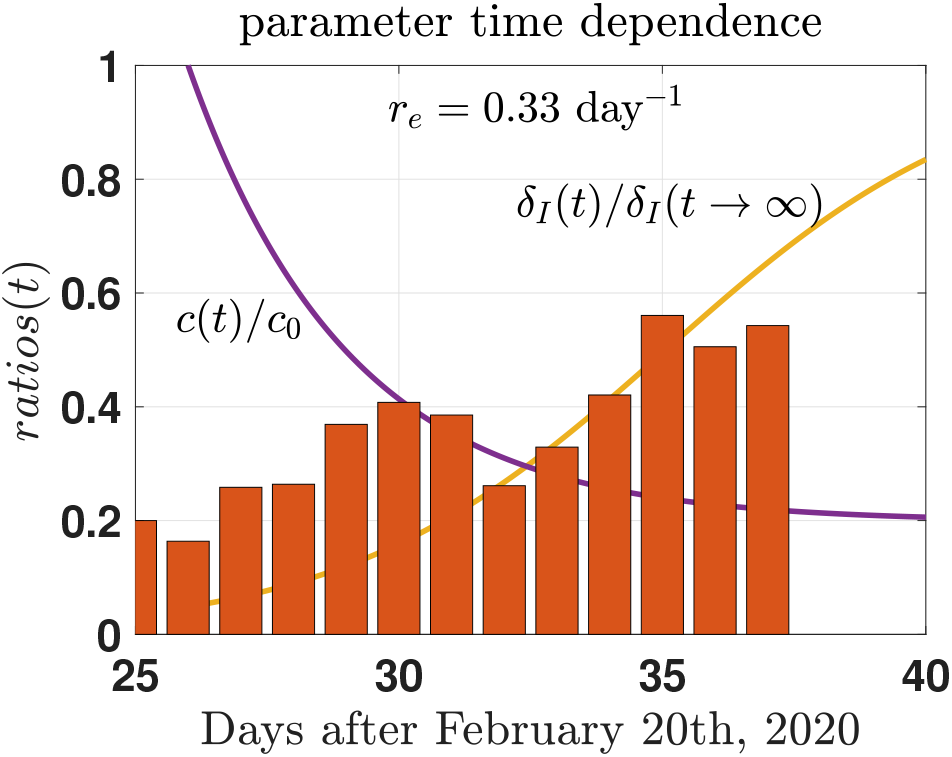
(color on line) Modelling the variation in time of the contact rate per person *c*(*t*) (*c*(*t* = 0) = *c*_0_) and of the rate from infected to quarantined classes *δ*_*I*_ (*t*) (cfr. Eqs.(9),(10)). The increment of the number of tests performed on the population is also graphically shown in the lower part of the figure to illustrate the consistence of the parametrization chosen. Data from refs. [14, 15].

The parametrization (10), in particular, is chosen to simulate the behavior of the screening system to select infected (and a-symptomatic) individuals. In Fig. 3 (upper panel), the per-day number of tests performed in Italy in the last weeks are explicitly shown. The number increases in time following the adopted measures, an enhancement coherent with the parametrization (10) as already emphasized in the caption of Fig. 2. On the other hand if one compares the behavior of the number of tests as shown in Fig. 3 (upper panel) with the percentage of positive tests (lower panel), one can conclude that the percentage is stable (around 22%) while the number of tests increases: a clear signal of the increased rapidity of the screening results.

**FIG 3.**
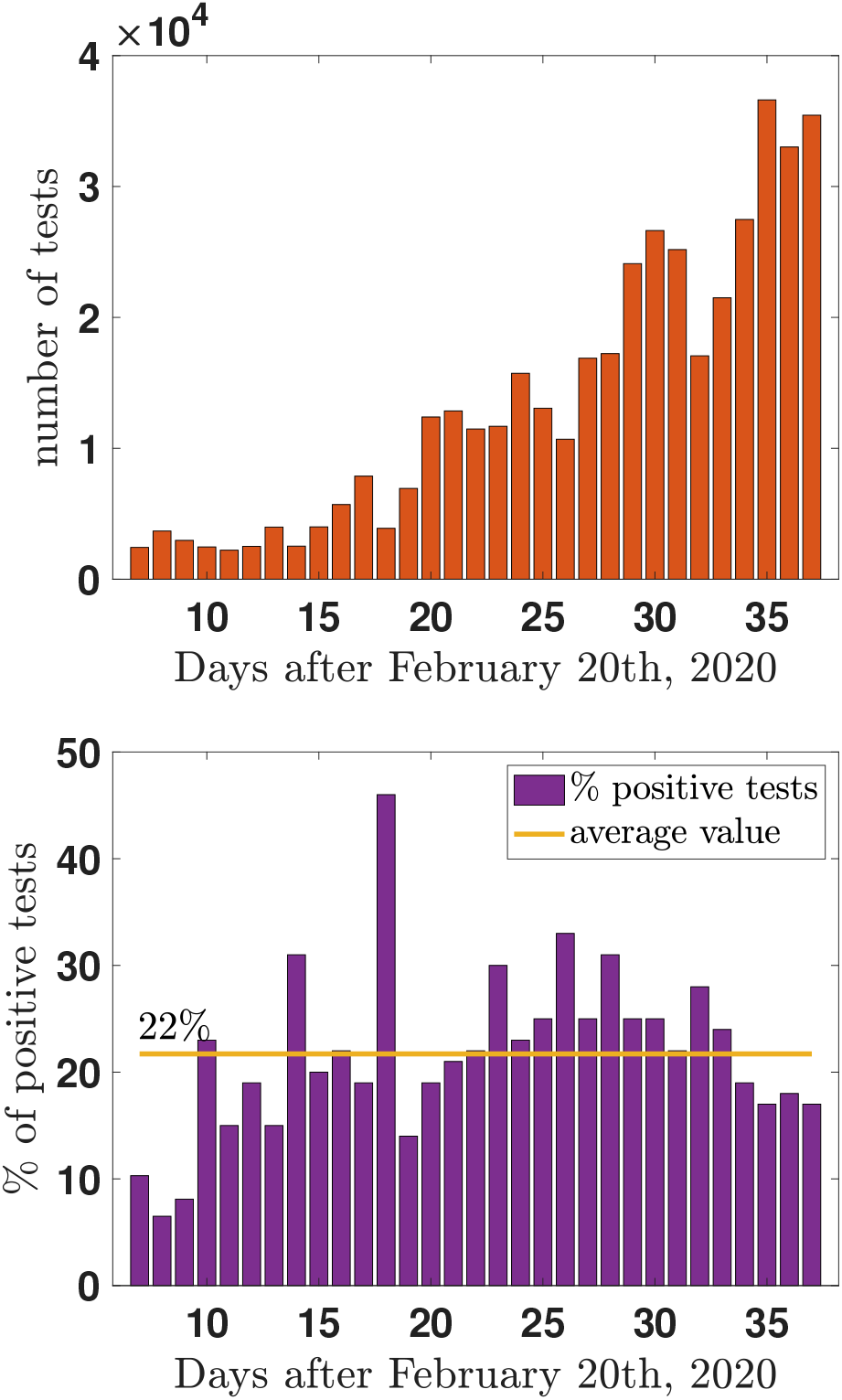
(color on line) **upper panel:** The per-day number of tests performed in Italy in the last weeks. **lower panel:** Percentage of positive test per day. The average value is also shown (continuous horizontal line). Data from refs. [14, 15].

### B. The effective (daily) Reproduction number

The next generation matrix (cfr. refs. [16, 17] has been used in refs. [8, 9] to derive an expression for the effective (daily) reproduction number which includes parametrization of the control measures. One has:

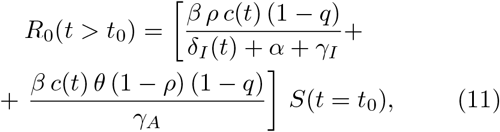

and it depends on time if the contact rate coefficient *c* and the quarantine rate *δ*_*I*_ are time dependent.

## III. RESULTS

### A. Modelling the early days of the outbreak

The present Section is devoted to studying the development of the epidemic event in Italy and the relevant parameters useful to mitigate the effects.

One can preliminarily look at the general behavior of the Infected compartment numbers, as shown In Fig. 4.

**FIG 4.**
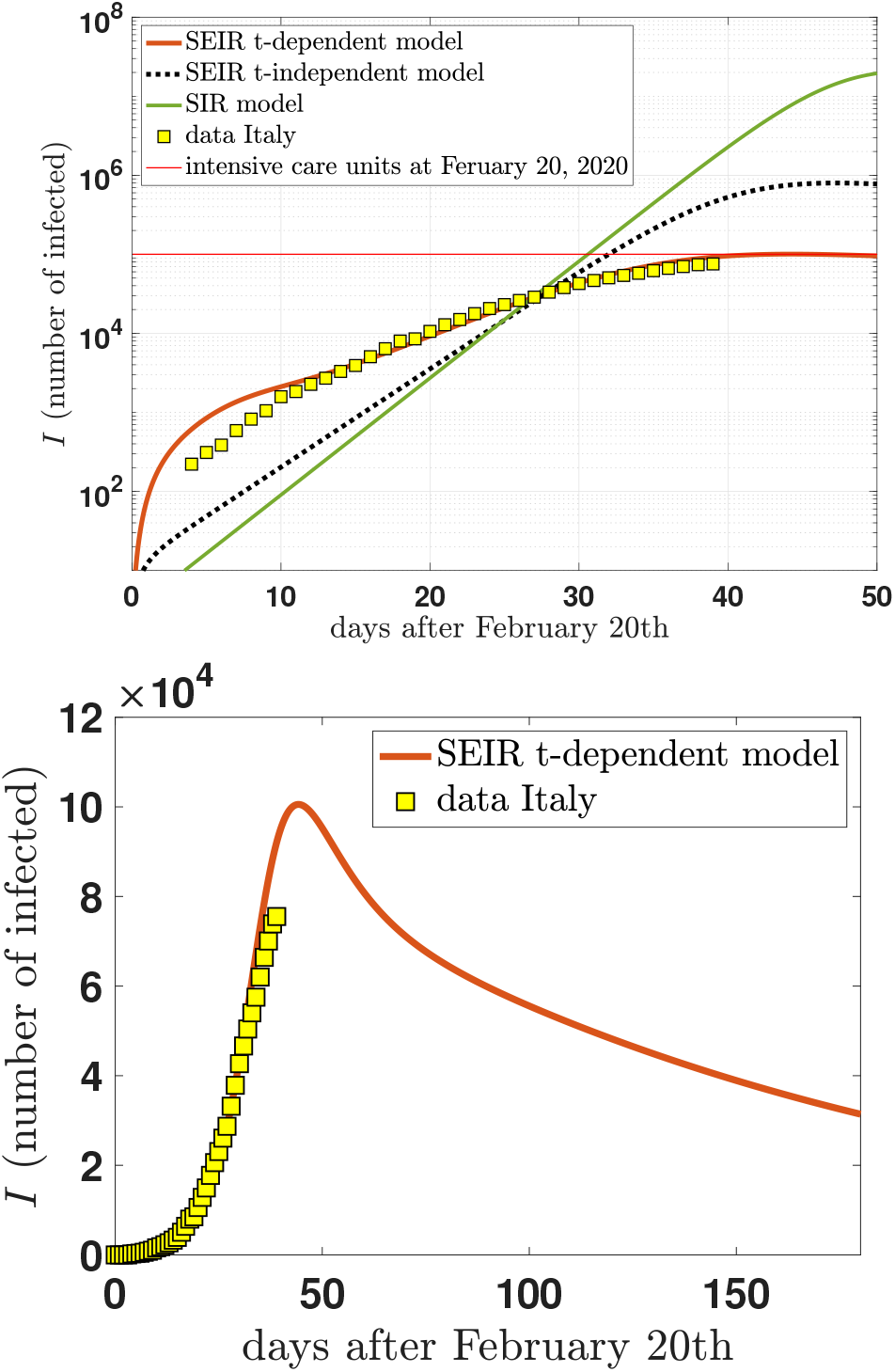
(color on line) **upper panel:** The daily Infected population in Italy as a function of the number of days after February 20 (day= 0) in a logarithmic scale. The yellow squares show the Italian data of the infectious population. The results take into account the variation in time of the measures and diagnose. In particular the decrease of the contact rate due to lack of contact for people solicited to remain home (see Eq. (9)) and the shortening of the diagnose tests (from 7 days to 8 hours (full curve), see also Eq. 10. The results are compared with a simple (time independent) *SEIR*-model including isolation and quarantine [8, 10] (dotted line) and a basic *SIR*-model predictions (*R*_0_ = 3.4). Data from refs. [14, 15]. **lower panel:** The results, of the time dependent model in linear scale, for the Infected population are compared with data.

The upper panel is on a logarithmic scale in order to fit different models. In particular, the simple SIR model (continuous green curve, *R*_0_ = 3.4) largely exceeds, in the hot period of the peak, the number of infected whose 5% needs intensive care (a rough estimate following the experience in China). The t-independent SEIR model containing all the stratified compartment as described by the Eqs. (1)-(8), largely lowers the peak value (cfr. [10] without reproducing the general behavior of the data (yellow squares), while the introduction of time dependence gives a prediction rather close to the observed cases. The linear curve (lower panel) only specifies the details and can be useful for further reference.

Let us now enter the discussion of the reproduction number. We investigate different scenarios in order to understand the effects of possible mitigation measures. We make reference to Fig. 5.

**FIG 5.**
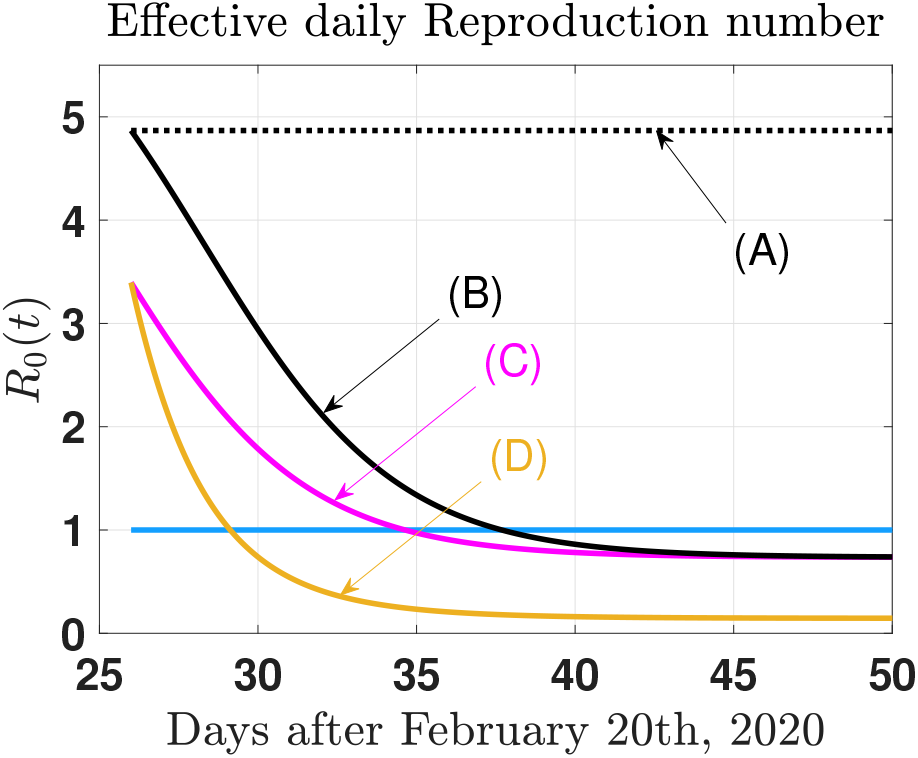
(color on line) The effective daily reproduction number as a function of time from Eq.(11) and within the different scenarios (A), (B), and (C) discussed in Section III. (A): SEIR time-independent model, significant and constant *R*_0_ *≈* 4.9. (B): SEIR time-dependent model. Reduced contact rate according to distancing measures; *c* = *c*_0_ = 2 day^*−*1^. Diagnostic time *t*_*I*_ reduced from 7 days to 8 hours. (C): SEIR time-dependent model. Reduced contact rate according to distancing measures; *c* = *c*_0_ = 2 day^*−*1^. Diagnostic time *t*_*I*_ reduced from 3 days to 8 hours. (D): (cfr. Section IV) SEIR time-dependent model with combined effects of the reduction of contact rate (9) and screening diagnostic time (10).

A. Scenario (A) refers to the time-independent SEIR model (cfr. upper panel of Fig. 4. The effective reproduction number is constant (cfr. Fig. 5). With the parameters of table I fitted on the Wuhan outbreak [8] and renormalized to reproduce the early data in Italy [10], one obtains *R*_0_*≈* 4.9. The value of *c* = *c*_0_ = 2 day^*−*1^ is rather low compared with the Wuhan rate (*c*_0_ *≈* 15 day^*−*1^) reflecting the different homogeneities of the two countries and the different social organization. It is shaped on the limited number of contacts after the control measures. The significant value of the reproduction number results in a massive outbreak hardly mitigated by isolation and quarantine. In particular, the model assumes a rate of infected to the quarantine of 7 days, a considerable interval of time before isolating symptomatic individuals. A hard consequence is related to the relatively low number of intensive care units available, as indicated in the same Figure by the red horizontal line.
B. Scenario (B) takes into account the time reduction of the screening test as parametrized in Eq.(10) in coherence with the test analysis of Section II A. In scenario (B), the time reduction is from 7 days to 8 hours, while the contact rate remains fixed and as low as *c* = 2 day^*−*1^. The net result is that the reproduction number can have values lower than 1, switching the outbreak off. The limiting values are 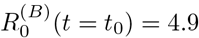 and 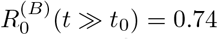. Fig. 5 illustrates the scenario as described in the caption.
C. Scenario (C) is analogous to scenario (B) with *c* = 2 day^*−*1^ and time reduction of the screening procedures from 3 days to 8 hours. Also in this case the reproduction number can have values lower than 1 and the limiting values are 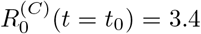 and 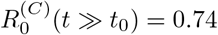. Fig. 5 illustrates the Scenario as described in the caption.

### B. Modelling the near future: secondary effects

Italy has been the first western country involved in the epidemic outbreak SARS-CoV-2. After 39 days (March 30th) the evolution is still a first expanding phase. However, after almost 20 days of isolation, the perspective of relaxing, at least partially, the social distancing measures is appearing in many discussions at different levels.

This Section is devoted to the quantitative study of a possible secondary event.

In this case, the time-dependent model can be used as a parametrization of the infected behavior and as a framework to estimate secondary effects.

Let us assume that at a given day (as a first hypothesis we fixed May 1st, 2020), the containment measures are totally or partially relaxed. The time evolution of the contact rate could be described as in Fig. 6. The isolation value, as discussed in the previous Section, assumes the limiting value *c ≈* 0.2 (isolation), and at day = 70 (from February 20th), the isolation is interrupted and the normalcy activated. Two scenarios are introduced: a full return to the previous style of life (rather unlike, but it represents a reference point) and a more realistic scenario with half isolation.

**FIG 6.**
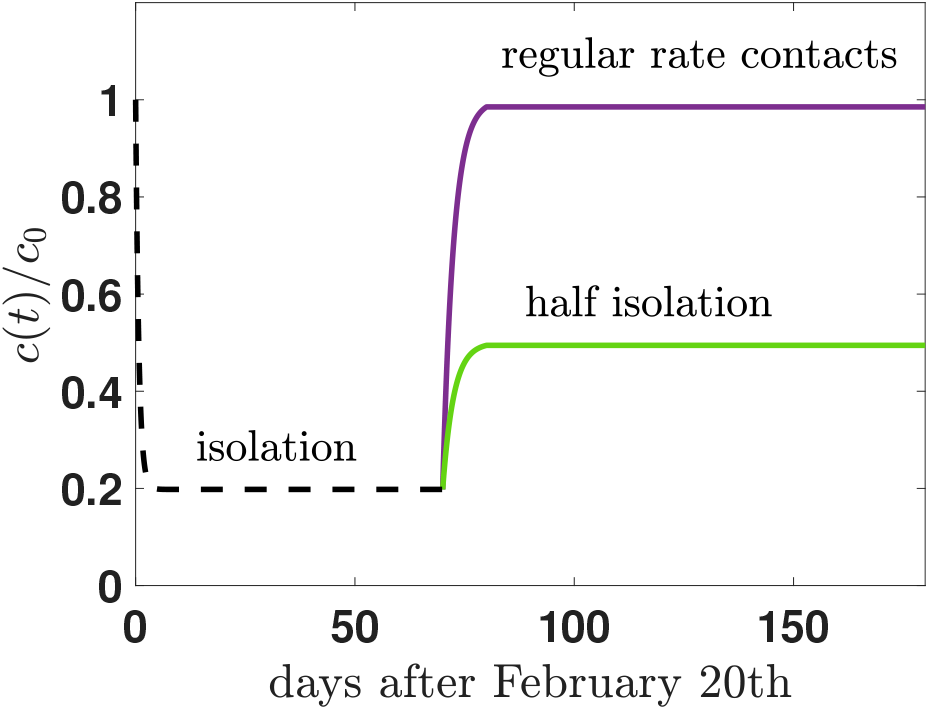
Time behavior of the contact rate simulating a possible secondary event in Italy at day=70, after February 20th, when the stringent measures of isolation are hypothetically relaxed. Two scenarios are introduced, the scenario of a rapid return to the old style of life (regular rate contacts), and a more realistic scenario where only half of the regular contacts are activated.

Assuming the parametrization of table I (see Appendix) and *θ* = 0.9 as the contribution of asymptomatic infected to the spread of the disease, one can easily fit the present behavior of the Infected compartment members. (cfr. Fig. 7, upper panel, dashed curve). The good comparison between data and model can be valid for a short time, but we are not interested in reproducing the exact numbers, but the relative effects of a possible secondary event. The occurrence of a subsequent event is unavoidable since the first outbreak did not exhaust its virulence: when normalcy is activated, a second peak appears, enhancing the tail value of the distribution.

**FIG 7.**
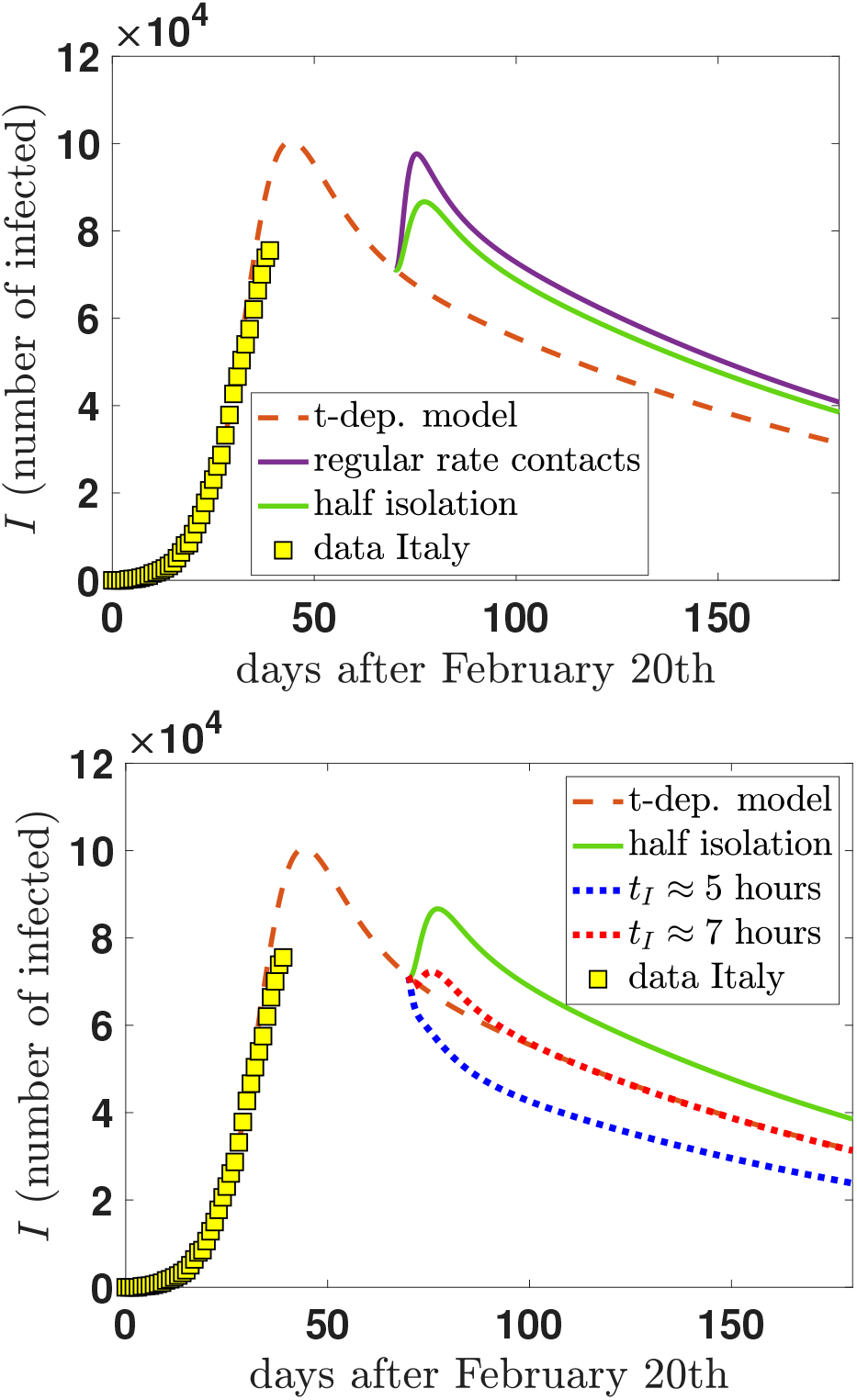
(color on line) **upper panel:** The time dependent model used to fit the data of infected in Italy (refs. [14, 15]) (dashed line) is then utilized to obtain a simulation of the reaction of the system to a reduction of the distancing measures at day = 70 and within the two different scenarios of Fig. 6. Strong secondary events are present in both cases. **lower panel:** If, at day = 70, half isolation measures are activated and, at the same time, the rapidity (*t*_*I*_) of the diagnostic tests is reduced from *t*_*I*_*≈* 8 hours (dashed line), one can substantially mitigate the effect of the secondary peak (continuous line). As examples: the red dotted line assumes a reduction from 8 to *≈* 7 hours, the blue dotted line from 8 hours to *≈* 5 hours.

Guided by the analysis of the effects of the technological developments in the screening phase done in the previous sections, one can try to see their effect again in the present secondary event, and the results of such an investigation are shown in the lower part of Fig. 7. Leaving the model evolving till day = 70 (May 1st) within the most strict measures (isolation), the same measures producing the best fit. May 1st one relaxes (partially or totally) the isolation control, the model will evolve following the new initial conditions fixed at that date. The peak of a new (secondary) outbreak appears, and its maximum value and duration depend on the rest of the parametrized conditions. Particularly relevant, once again, the value of *t*_*I*_ = 1*/δ*_*I*_ reached during the transition time to normalcy. Lowering the diagnostic time to *t*_*I*_*≈* 7 hours, or *t*_*I*_ *≈* 5 hours has the power to substantially mitigate the secondary events, in particular, if one keeps “half isolation” (as schematically designed in Fig. 6).

## IV. DISCUSSION

Our results on the numerical values of the (effective daily) reproduction number *R*_0_ show that a social distancing of 2 contacts per day is not adequate to reduce its value below 1.7, even with a test rapidity *t*_*I*_ = 1 day to quarantine infected people. Our study suggests that the two parameters have to be activated at the same time. Requesting strict social distancing without reaching a critical value for *R*_0_ in a reasonable number of days risks failure and the loss of faith in the adopted measures. The most favorable Scenarios (D) and (E) (cfr. lower panel of Fig. 4) need from 10 to 15 days before reaching values of *R*_0_ *<* 1 despite a screening rapidity of 8 hours (from a starting point of 3 days). The inertia of the distribution can force the time interval up to one month before seeing visible effects. The improvement of rapid diagnosis of SARS-CoV-2 with automation and a large number of sample processing is essential in order to implement infection prevention measures lowering *R*_0_ significantly.

After March 8th, the measures proposed to increase the social distancing in Italy have been enhanced each week. This escalation of rules intended to lower the contact rate (already as low as 2 contacts per day, as assumed in the scenarios of Section III. The combined effect of the imposed isolation and a strong reduction of screening time can be seen in the lower curve of Fig. 5 (scenario (D)). The cumulative effect reduces the delay from 10 days to 5 days to reach *R*_0_*≈*1 and, in a further 5 days, *R*_0_ *≈* 0.15, a value drastically low to see, day by day a 30% reduction of newly infected people each day. The corresponding parameters are *c≈* 1 contact per day and *t*_*I*_ *≈* 10 hours.

These results seem to be entirely consistent with the Comment by Anderson *et al*. [12].

The flexibility of the model we are using can allow the introduction of the effects of such further control restrictions. Eq. (9) shows how such a measures have been further implemented reducing the contact rate from *c*(*t* = *t*_0_) = *c*_0_ = 2 day^*−*1^ to *c*(*t»t*_0_) *≈* 0.4. In this case the reproduction number assumes the limiting values 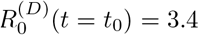 and 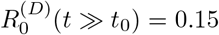. Figs. 5 shows graphically the outcomes. The time interval to reduce the reproduction number to be less than 1 is drastically lowered and (probably) one can even see such an effect already at the early stage of the epidemic looking at the recent data on upper panel of Fig. 5 (yellow squares).

The same approach can be applied to evaluate the rise of a secondary epidemic event at the moment of a partial relaxation of the isolation measures adopted. Our findings manifest an unavoidable occurrence of subsequent events. However, they can find our countries prepared thanks to the delay obtained in the first phase through isolation. The strategy to implement is based mainly on technological resources opening a new era of fast screening. For a more in-depth discussion of the results, one wonders if the choice of the moment can substantially influence the subsequent event. The need to save intense care departments from a flood of hospitalizations pushes the inertia of the event and produces a long time tail; it is tough to keep isolation for a corresponding period. However, one can analyze the effects of the shift in time. In Fig. 8, the results of this further analysis. The technological improvement of the screening process remains the essential mitigation ingredient, and the structure of the subsequent event does not change enough to suggest a further delay.

**FIG 8.**
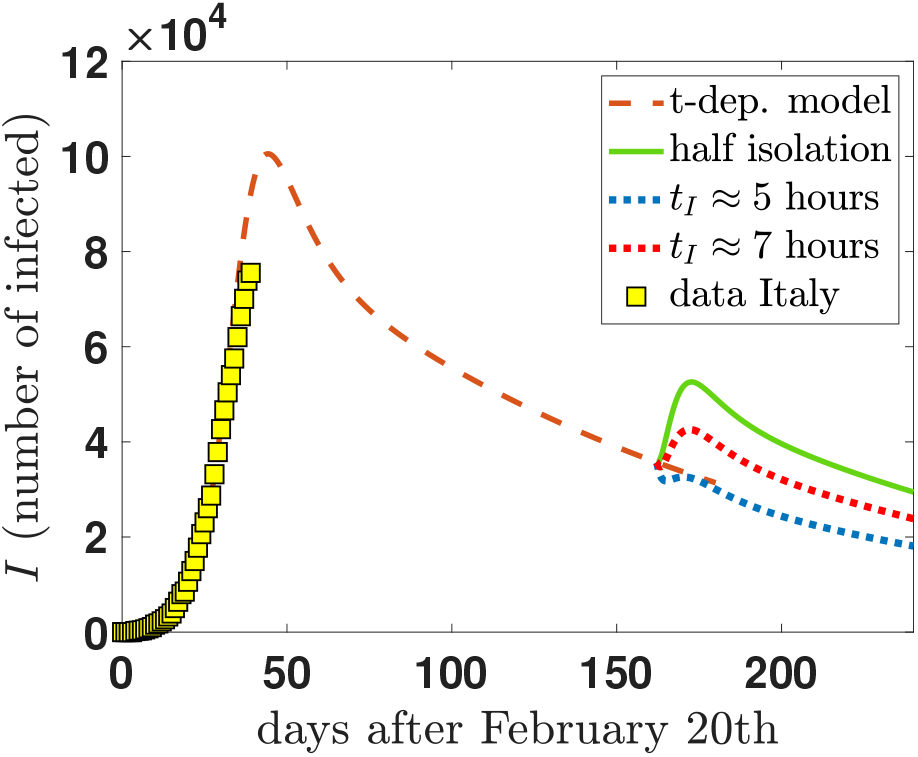
(color on line) If at day = 162 (August 1st), half isolation measures are activated and, at the same time, the rapidity (*t*_*I*_) of the diagnostic tests is reduced from *t*_*I*_ *≈* 9 hours (dashed line), one can substantially mitigate the effect of the secondary peak (continuous line). As examples: the red dotted line assumes a reduction from 9 to *≈* 7 hours, the blue dotted line from 9 hours to *≈* 5 hours.

### a final comment

Italy is the first European country massively experiencing the pandemic SARS-CoV-2 event. Finding adequate measures against the invasion of a new (and dangerous) virus is requesting an extraordinary and epochal effort to generate social measures rather unknown and able to profoundly modify the social behavior, both at the personal and collective levels. The single person is experiencing an entirely new horizon, and the death of a portion of aged people is cutting the only link with experience able to react with a verified, effective way. The testimonial given by the medical personnel is an example that such a link has been fructose. The present work has been motivated by the (even remote) possibility that the scientific approach to the pandemic event can help to assume the most efficient measures.

A pandemic event can only be stopped (mathematically) only by a large number of unsusceptible people. Waiting for a vaccine able to increase the number of un-susceptible, the generic arms we have are restricted to isolation and quarantine. However, the present study offers another evidence that our civilization’s highest technological level can speed up the population screening process to identify the individuals to put in isolation. The consideration that the contact rate cannot be reduced under a minimum level is contrasted efficiently by the drastic reduction of the time *t*_*I*_ needed for an isolation decision.

## V. JUNE 2020: A LONGTERM VIEW

With the kind permission of the editor and referees, let us include a long term analysis of the model proposed in the previous sections. In practice, one would like to answer a simple question: how does the model describe data in the long term? To answer, the semi-quantitative analysis proposed needs a better implementation, adding a quantitative investigation of the parameters of the model when compared with the data of the outbreak in Italy.

To this aim one can make use of a more detailed investigation [18] based again on the model of Eqs. (1)-(8), and Fig. 1.

The significant advantage is the knowledge of the statistical uncertainties on the values of the model parameters, uncertainties which have a methodological, theoretical origin in the procedure of the data-fit. The Markov Chain Monte Carlo (MCMC) method is used to fit the model, and an adaptive MetropolisHastings (M-H) algorithm is adopted to carry out the MCMC procedure. The algorithm is used for four concatenated runs with the following number of iterations: 100.000-50.000-25.000-10.000.The procedure runs in Matlab with the MCMC toolbox for Matlab.

For the model to be as predictive as possible, we applied it to two different geographical regions: Italy itself and a region of Italy, Lombardy, where the epidemic event is manifesting in a specific and violent way. Italy is a large geographical area, and the data are averaged on quite different realizations of the same epidemic event in various regions from north to south. Lombardy is a much more homogeneous region, and it can be considered even more representative of the epidemic progression without excessive geographical and social smearing.

### A. Long term analysis: Italy

In Fig. 9, a first set of results comparing model predictions and data in Italy. In particular, the lower curves compare the daily reported case in Italy after February 24th and the model predictions, which include the statistical uncertainties. The green part of the data refers to the data used to fix the parameters and not represent predictions. The yellow data show the pure predictive power of the present approach. The number of days where the data remain within the theoretically predicted area is of the same order of magnitude of the number of days used for the fit (*∼* 40 days). The model does not describe the last part of the data. Of course, one must keep in mind the number of social changes (and restriction or relaxation measures) present in the long term, changes that are not necessarily included in the model parametrized within the first period of the outbreak. Besides, as already said, the lack of homogeneity between the different areas of the nation makes the modelling quite hard as far as quantitative average predictions are concerned.

**FIG 9.**
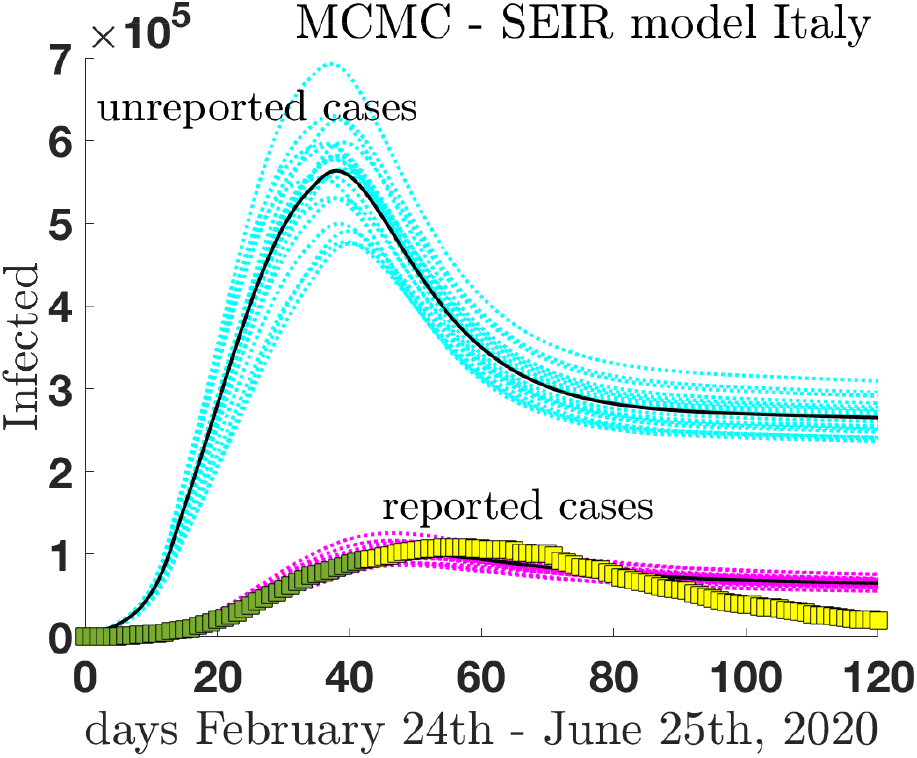
(color on line) A MCMC analysis of the infected individuals (reported cases) in Italy as a function of time. The data for infected individuals of refs. [14, 15] are compared with the SEIR model predictions which include statistical uncertainties (lower curves). The green data refer to the data set used to fix the model parameters, while the yellow set spans the purely predicted period. To complete the analysis the reported cases are compared with the predictions for the class of infected non reported cases (asymptomatic) in the same period of time.

However, if the long-term comparison is quite demanding, the model has predictive power for other kinds of stratification, such as the distribution of unreported cases (asymptomatic). In the same of Fig 9, the data, and predictions for the reported cases are compared with the predictions for the unreported cases, including the class of asymptomatic. The total number of infected individuals receive a substantial contribution from the distribution of unreported cases.

In Fig. 10, the comparison between the model predictions and the data is even more stringent. The data of the daily *new* (reported) cases, from February 24th on, are compared with the predictions of the SEIR model. This kind of comparison involves the variations of the daily reported cases, a particularly demanding test for the model. The data show a large dispersion due to the influence of strategic decisions after the lockdown period (i.e. after May 4th). This influence explains the data around May 6th, when a significant fraction of infected isolated individuals was reported as recovered in a region (Lombardy) with the largest number of reported infected cases in Italy. The behavior of the data follows rather consistently the model predictions of the reported cases of Eq. (7) once scenarios for the mitigation of the lockdown are introduced. In particular, the data seem to follow rather consistently a scenario (green lines) [18] where the contact rate excludes a large part of social events (school, sports, large events) and it includes the introduction of tracing and quarantine. The fact that the longterm data (after May 4th) are well reproduced by the model means the distancing and tracing measures proposed in Italy have a positive impact captured by the SEIR model. More dramatic scenarios (as for described, for instance, by the cyan curves) have been avoided.

**FIG 10.**
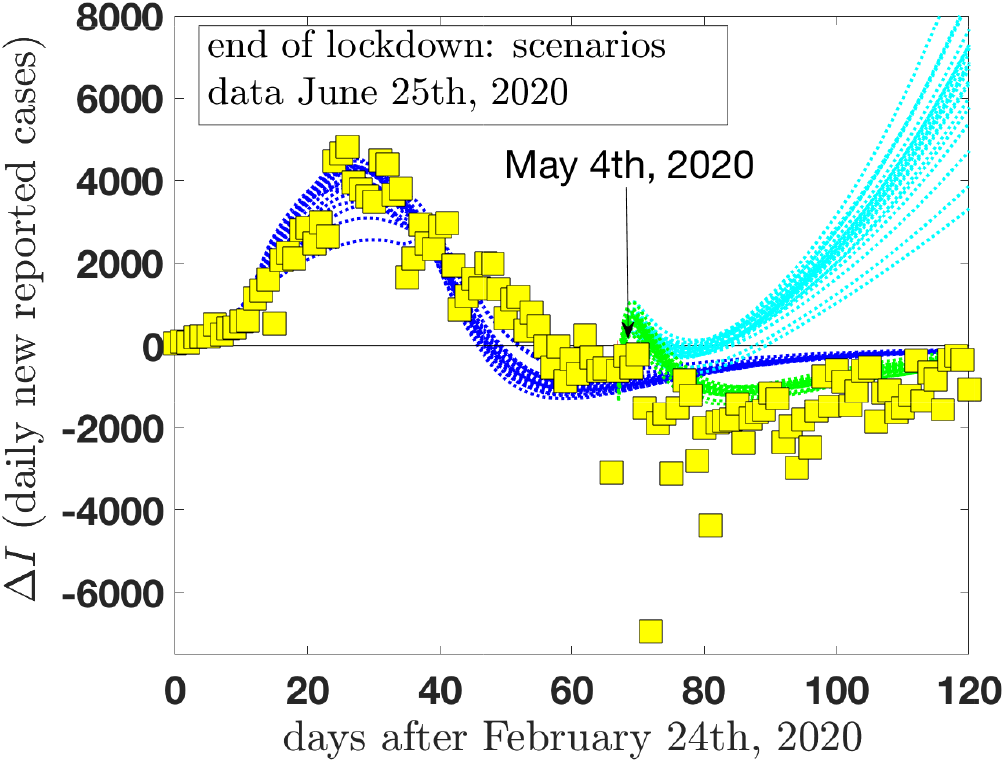
(color on line) The model predictions for the daily new reported cases after February 24th, in Italy, are compared with the official data of refs. [14, 15]. In addition to the fully locked scenario (blue lines), two other scenarios are included for a mitigation of the lockdown period in Italy starting from May 4th. Scenario A (cyan lines): a social contact rate somewhat similar to the regular rate (only a reduction of a factor of two is included) would result in a violent secondary outbreak. Scenario B (green lines): A stronger reduction of the contact rate which excludes a large part of social events (school, sports, large events…) with the further inclusion of tracing and quarantine.

### B. Long term analysis: Lombardy

The second analysis of longterm predictions refers to the Lombardy region. A quantitative comparison is shown in Fig. 11. The lower curves compare the reported cases in Lombardy, analogous to the results of Fig. 9 for Italy. As anticipated, the results show a somewhat consistent agreement between the model predictions and the data. This agreement is due to the larger homogeneity of the geographical and social characteristics of the area. The evident discontinuity around May 6th shows the effect of strategic decisions after the lockdown period, as already discussed in the figure caption of Fig. 10. To complete the analysis, the same Fig. 11 reports (in analogy with Fig. 9) also the predictions for the unreported (asymptomatic) cases in Lombardy region..

**FIG 11.**
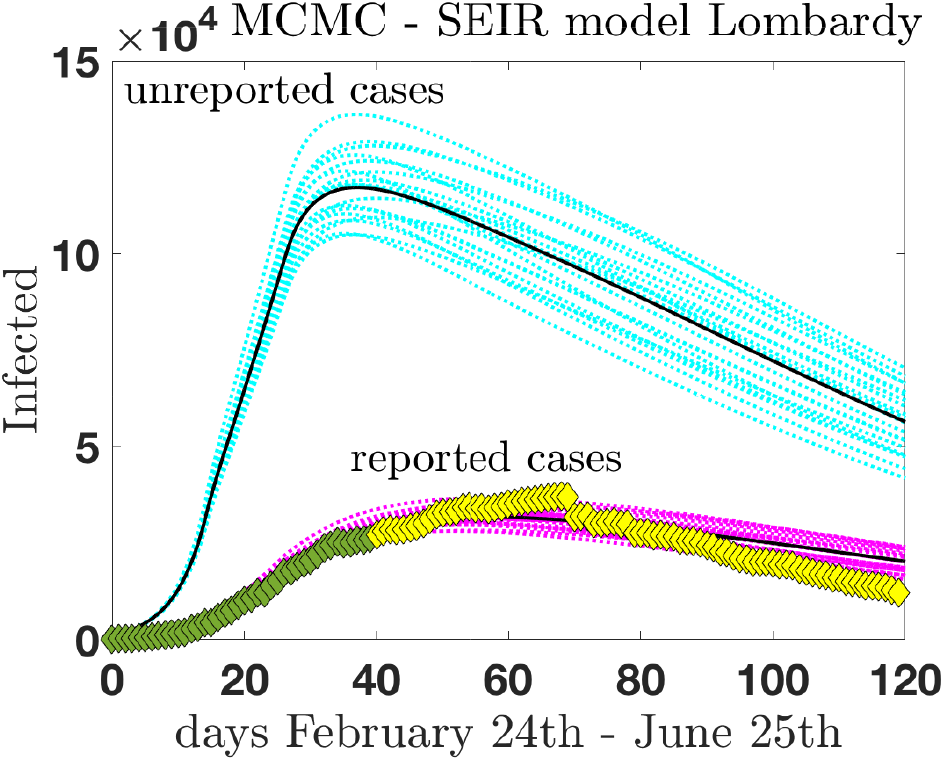
(color on line) A MCMC analysis of the infected individuals (reported cases) in Lombardy y as a function of time. The data for infected individuals of refs. [14, 15] are compared with the SEIR model predictions which include statistical uncertainties (lower curves). The green data refer to the data set used to fix the model parameters, while the yellow set spans the purely predicted period. To complete the analysis the reported cases are compared with the predictions for the class of infected non reported (asymptomatic) cases in the same period of time.

### C. to conclude

The Section June 2020: a longterm view discusses the comparison between model predictions and actual data set of the SARS-CoV-2 event in Italy as asked by the referee and the editor. The section is rather informative and required the use of a refined statistical model [18], always based on the Eqs. (1)-(8), and Fig. 1. The comparison shows the goodness and weakness of the model keeping in mind the number of social changes (and restriction or relaxation measures) present in the long term. Besides, the lack of homogeneity between the different areas of the nation makes the modelling quite hard for quantitative long-term average predictions. To better understand the actual power prediction of the model, a more homogeneous sub-area has been discussed (Lombardy), where the virulence of the epidemic event has been particularly high. In that case, the comparison shows to be even more favorable.

## Data Availability

all data are included in the official web sites
Ministero della Salute
Il Sole 24 ORE

http://www.salute.gov.it/portale/nuovocoronavirus/

https://lab24.ilsole24ore.com/coronavirus/

## Contributors

MCT and RF programmed the model. CC provided the data from online sources. GVD interpreted the study findings, contributed to the manuscript. All authors interpreted the findings, contributed to writing the manuscript, and approved the final version for publication.

## Acknowledgements

MCT thanks the members of the Institut de Physique Théorique, Université Paris-Saclay, for their warm hospitality during a recent visiting period. He also thanks to the Physics Department of Valencia University for support and friendly hospitality. Useful remarks by Hervé Tricoire, Paris University/CNRS, and critical reading of the manuscript by Cristina Di Girolami, Le Mans University, are gratefully acknowledged.

## Research in context

### Evidence before this study

We searched PubMed, BioRxiv, and MedRxiv for articles published in English from inception to February 20th, 2020, with the keywords 2019-nCoV, novel coronavirus, COVID-19, SARS-CoV-2 AND reproduction number Italy, Italy transmission. We found several estimates of the basic reproduction number (*R*_0_) of severe acute respiratory syndrome coronavirus 2 (SARS-CoV-2), including average exponential growth rate estimates based on inferred or observed cases at a specific time-point and early growth of the outbreak. However, we identified no estimates of how *R*_0_ had changed in Italy since control measures were introduced in March or estimates that jointly fitted data within Wuhan and Italy and no estimates of secondary effects in Italy.

### Added value of this study

In our study, we estimate how the transmission has varied over time in Italy, identify a decline in the reproduction number in the late period as a function of the introduction of large scale control measures, and show the potential implications of estimated secondary effects.

### Implications of all the available evidence

SARS-CoV-2 is currently showing sustained transmission in Italy, with a substantial risk of outbreaks in different areas. However, the implementation of rapid technologies during screening can largely influence the value of the reproduction number and support a (partial) relaxation of the isolation measures without introducing a substantial risk of secondary outbreaks.

## Appendix A: Parameters and tables

We summarize in the table I of the present appendix the parameters of the SEIR-type model as proposed by Tang *et al*. in ref. [8]. Table II is devoted to summarizing the initial conditions imposed on the SEIR solutions in the numerical calculation.

**TABLE I.**
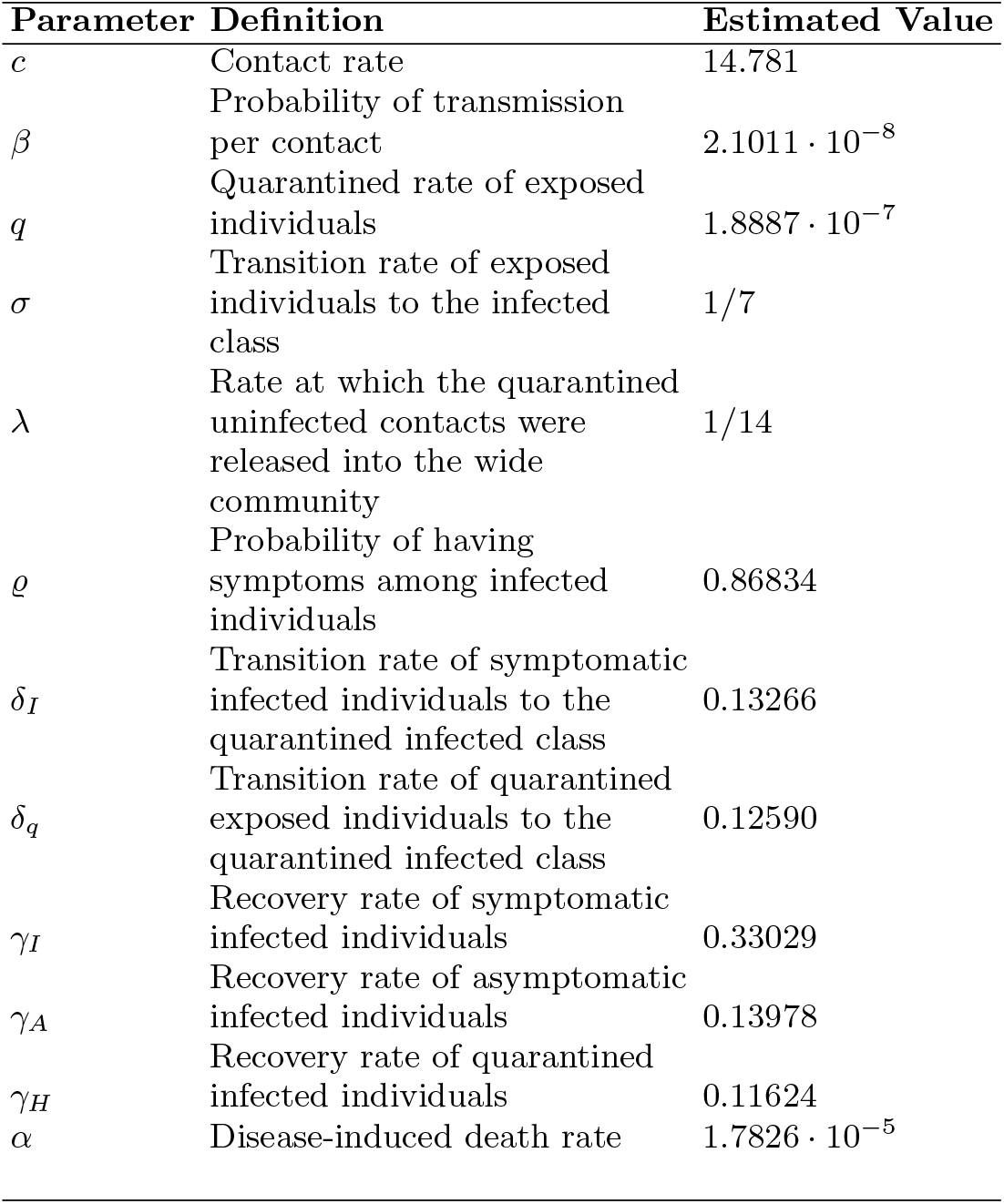
Parameters of the SEIR model description of the Wuhan outbreak.

| Parameter | Definition | Estimated Value |
| --- | --- | --- |
| $c$ | Contact rate | 14.781 |
| $\beta$ | Probability of transmission per contact | $2.1011 \cdot 10^{-8}$ |
| $q$ | Quarantined rate of exposed individuals | $1.8887 \cdot 10^{-7}$ |
| $\sigma$ | Transition rate of exposed individuals to the infected class | 1/7 |
| $\lambda$ | Rate at which the quarantined uninfected contacts were released into the wide community | 1/14 |
| $\varrho$ | Probability of having symptoms among infected individuals | 0.86834 |
| $\delta_I$ | Transition rate of symptomatic infected individuals to the quarantined infected class | 0.13266 |
| $\delta_q$ | Transition rate of quarantined exposed individuals to the quarantined infected class | 0.12590 |
| $\gamma_I$ | Recovery rate of symptomatic infected individuals | 0.33029 |
| $\gamma_A$ | Recovery rate of asymptomatic infected individuals | 0.13978 |
| $\gamma_H$ | Recovery rate of quarantined infected individuals | 0.11624 |
| $\alpha$ | Disease-induced death rate | $1.7826 \cdot 10^{-5}$ |

**TABLE II.** default

| Initial values | Italy | Wuhan |
| --- | --- | --- |
| $N$ | $6 \cdot 10^7$ | $1.1 \cdot 10^7$ |
| $E(0)$ | 100 | 105 |
| $I(0)$ | 3 | 28 |
| $A(0)$ | 100 | 53 |
| $S_q(0)$ | 0 | 739 |
| $E_q(0)$ | 0 | 1 |
| $H(0)$ | 0 | 1 |
| $R(0)$ | 0 | 2 |

